# Clinical validation of novel second-trimester preterm preeclampsia risk predictors combining clinical variables and serum protein biomarkers

**DOI:** 10.1101/2022.12.21.22282936

**Authors:** Louise C. Laurent, George R. Saade, Glenn R. Markenson, Kent D. Heyborne, Corina N. Schoen, Jason K. Baxter, Sherri A. Longo, Leonardo M. Pereira, Emily J. Su, Matthew K. Hoffman, Garrett K. Lam, Angela C. Fox, Ashoka D. Polpitiya, Md. Bahadur Badsha, Tracey C. Fleischer, Thomas J. Garite, J. Jay Boniface, Paul E. Kearney

## Abstract

**Objective:** To develop and validate mid-trimester preterm preeclampsia (PE) risk predictors combining clinical factors and serum protein analytes, and to compare their performance with those of widely used clinical and risk assessment algorithms endorsed by professional societies.

**Methods:** This was a secondary analysis of data from two large, multicenter studies in pregnant individuals (PAPR, NCT01371019; TREETOP, NCT02787213), originally conducted to discover, verify, and validate a serum proteomic predictor of preterm birth risk. Serum protein abundances were determined by mass spectrometry. Classifier models combined one or two novel protein ratio(s) with a composite clinical variable, denoted as ClinRisk3, which included prior PE, pre-existing hypertension, or pregestational diabetes. Predictive performance was assessed for the full validation cohort and for a subset that had early gestational age (GA) dating via ultrasound. Classifier performance was compared directly to the U.S. Preventive Services Task Force (USPSTF) algorithm for identification of pregnancies that should receive low-dose aspirin (LDASA) for PE prevention.

**Results:** Nine of nine prespecified classifier models were validated for risk of preterm PE with delivery <37 weeks’ gestation. Areas under the receiver operating characteristic curve ranged from 0.72-0.78 in the full validation cohort, compared to 0.68 for both ClinRisk3 alone and for the USPSTF algorithm. In the early GA dating subcohort, an exemplar predictor, ClinRisk3 + inhibin subunit beta C chain/sex hormone binding globulin (ClinRisk3+INHBC/SHBG) showed a markedly lower screen positive rate (11.1% vs 43.3%) and higher positive predictive value (13.0% vs 5.0%) and odds ratio (9.93 vs 5.24) than USPSTF. Its performance was similar in nulliparas and all parities.

**Conclusion:** Nine preterm PE risk predictors were identified, validated in an independent cohort, and shown to be more predictive than the USPSTF-endorsed algorithm. Our results indicate that a single blood test performed in the first half of pregnancy can be used for personalized PE risk assessment, particularly for pregnancies with minimal or no identified clinical risk factors, including nulliparas. Results can be used to guide personalized pregnancy management, including but not restricted to LDASA for PE prophylaxis, and serve as a basis for developing new prevention strategies.

**Funding Source:** Sera Prognostics, Inc.

**Précis:** Combining novel serum protein biomarkers and selected clinical variables for preterm preeclampsia prediction outperforms widely used clinical risk assessment algorithms currently recommended by practice guidelines.

## Introduction

Preeclampsia (PE) occurs in ∼5-7% of pregnancies^1^ and is implicated in 14% of maternal^2^ and 10-25% of perinatal^3^ deaths. It is associated with approximately one-quarter of all preterm births (PTBs) and 50% of medically indicated PTBs.^4^ The only cure for PE is delivery, with other therapies focused on preventing or delaying onset (low-dose aspirin [LDASA]),^5^ preventing end-organ damage (antihypertensive medications to prevent strokes^6^ and magnesium sulfate to prevent seizures),^7^ and reducing neonatal complications of prematurity (antenatal steroids).^7^ Because PE-associated perinatal morbidity and mortality and healthcare costs correlate with prematurity,^8,9^ accurate prediction of preterm PE (gestational age [GA] at delivery [GAD] <37 weeks) is of greater clinical and economic value than prediction of term PE.

Until recently, PE prediction relied largely on clinical risk factors, including: PE in a previous pregnancy; high body mass index (BMI); maternal chronic hypertension, diabetes, renal disease, and autoimmune conditions; and obstetric characteristics such as multiple gestation and use of assisted reproductive technologies.^7^ PE and gestational hypertension in a previous pregnancy are among the most predictive and commonly used risk factors but are irrelevant in nulliparous pregnancies.

Numerous PE-associated molecular biomarkers have been reported,^10–13^ including soluble fms-like tyrosine kinase 1 (sFlt-1) and placental growth factor (PlGF). In patients with clinically suspected PE, sFlt-1 and PlGF measurement enables earlier diagnosis and/or modestly decreases adverse maternal outcomes.^14,15^ but do not adequately predict PE risk in asymptomatic pregnancies. Strategies incorporating combinations of serum analytes, patient history, blood pressure, and uterine artery pulsatility index accurately predict PE risk in asymptomatic pregnancies but are not widely used, as they require sophisticated equipment with trained sonographers along with molecular analyses. ^16,17^ Also, sFlt-1 and PlGF clinical studies have focused on late-onset PE; included only cases conforming to classical definitions; or grouped PE cases across all GAs and/or severities together. Since PE is highly heterogeneous,^18,19^ biomarker studies should at minimum assess performance for early-onset and late-onset cases separately.^8,20,21^

There is an urgent need for an accurate, safe, and practical predictor for preterm PE, particularly for nulliparas. A predictor with good AUC (>0.70), sensitivity (>50%), and PPV (>5%) at a reasonably low screen positive rate (SPR) (<15%) would markedly improve identification of high-risk pregnancies for LDASA prophylaxis, increased surveillance, and research studies compared to current clinical paradigms, while a high negative predictive value (NPV; >99%) would enable low-risk patients to safely forgo or discontinue unnecessary interventions. This study aimed to develop and validate a mid-trimester predictor comprising molecular biomarkers and relevant clinical factors across demographically, geographically, and temporally diverse populations, comparing predictive performance to that of currently used risk assessment paradigms.

## Methods

Biomarker development was based on U.S. National Academy of Medicine guidelines.^22^ This was a secondary analysis, using data from two large, multicenter U.S. cohort studies, conducted to discover, verify, and validate risk predictors for preterm PE. The Proteomic Assessment of Preterm Risk (PAPR, NCT01371019)^23^ and the Multicenter Assessment of a Spontaneous Preterm Birth Risk Predictor (TREETOP, NCT02787213)^24^ studies were prospective, observational studies conducted to develop a maternal serum-based proteomic biomarker for PTB risk stratification. Study approvals were provided by site IRBs, and participants provided written informed consent.

PAPR and TREETOP enrolled gravidas ≥18 years of age with singleton pregnancies from 17^0/7^-28^6/7^ or 17^0/7^-21^6/7^ weeks’ gestation, respectively, and excluded pregnancies with planned delivery <37^0/7^ weeks’ gestation, major anomalies or chromosomal disorders, or planned cervical cerclage. GA was determined according to ACOG criteria^25,26^. In TREETOP, progesterone use after 13^6/7^ weeks’ gestation was an exclusion criterion. Demographic, medical and pregnancy history, medication, and post-delivery maternal and infant outcome information was collected. No attempt was made to control for LDASA use in these studies. Adjudication of PE and PTB cases was blinded, occurring prior to database lockdown and analyses.

Discovery was performed in a PAPR-derived cohort that was distinct from the TREETOP-derived verification and validation cohorts (**Fig. 1**). The discovery and verification/validation cohorts included blood samples drawn 18^0/7^-22^6/7^ and 18^0/7^-20^6/7^ weeks’ gestation, respectively. The discovery cohort used a broader GA at blood draw (GABD) window to increase sample size. TREETOP subjects enrolled before April 18, 2018 were randomly assigned by an independent third-party statistician to blinded verification (one-third) or validation (two-thirds) cohorts.^24^ The validation cohort also included six months of later enrollment at four additional sites not represented in the verification cohort. Cases were defined as subjects with PE who delivered <37 weeks’ gestation, and controls were defined as noncases, inclusive of term PE and sPTB.

**Fig. 1.**
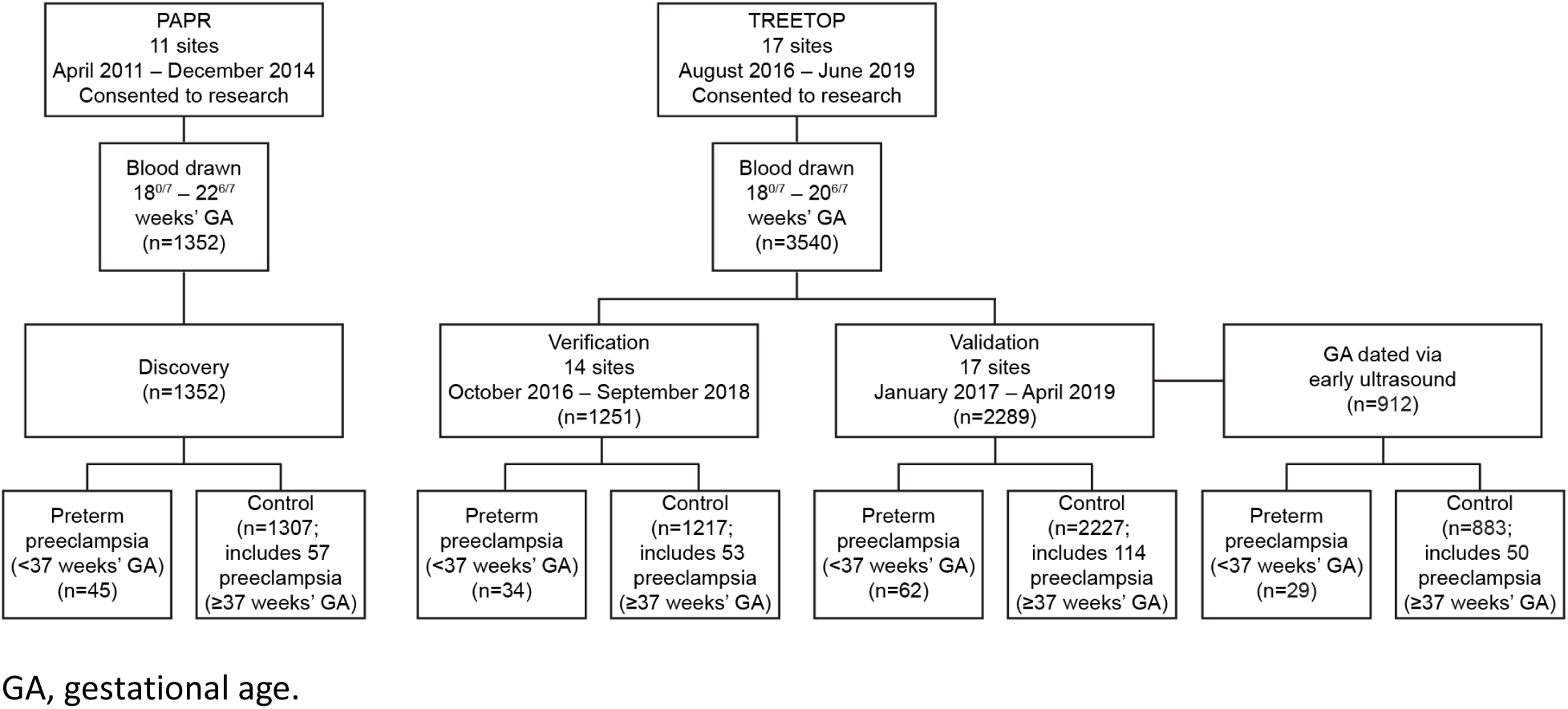
Distribution of subjects in the discovery, verification, and validation cohorts.

Maternal blood was collected, processed and stored as described previously.^23,24^ Serum samples were depleted of abundant proteins, reduced, alkylated, and digested with trypsin. Stable isotope standard (SIS) peptides were added post-digest, and peptides were analyzed by a liquid chromatography-multiple reaction monitoring mass spectrometry assay.^23,27^ Quantification was based on peptide response ratios, wherein the area of each endogenous peptide was divided by that of its corresponding SIS. Sample batches were required to meet quality control metrics.

Biomarker measurements were made for 77 proteins, comprising those enriched for pathways relevant to pregnancy conditions and quality control proteins. Proteins for building classifier models were prefiltered using analytical performance criteria, including mass spectrometry peak area cutoffs, coefficients of variance from replicate quality control samples, and preanalytic stability. Simple models were then constructed to avoid overfitting and to leverage protein ratios for normalizing and amplifying predictive signals^23^. Models included one novel two-protein ratio and a composite clinical variable. Based on strong predictive performance in nulliparous individuals, the log ratio of insulin-like growth factor-binding protein 4 (IBP4) to sex hormone-binding globulin (SHBG) was included in models as a second two-protein ratio unless SHBG was part of the novel ratio. Among many clinical factors specified in current practice guidelines,^7,28–32^ three (PE in a previous pregnancy, pre-existing hypertension, and pregestational diabetes) were selected and combined into one variable (ClinRisk3), which was deemed positive if any was true for the subject. This simple, fixed approach reduced overfitting, leveraged prior biomarker knowledge, and produced classifiers that followed two model structures:

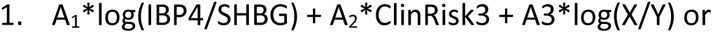

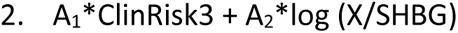

where X and Y are protein analytes, identified herein by Uniprot name without species identifier. Model coefficients A_i_ were fit using logistic regression.

In discovery, models were ranked by a combination of area under the receiver operating characteristic curve (AUC) and correlation with GAD amongst preeclamptic subjects. Candidate models ascertained in discovery were confirmed and tuned in verification to account for temporal changes in care and population, then validated by an independent third-party statistician. Each classifier was initially validated using AUC, then *P*-value, for three score thresholds distinguishing higher from lower risk. Thresholds were predefined based on discovery performance to allow for sensitivity and specificity tradeoffs and to enable use in varied clinical settings.

Previously we showed that for prediction of spontaneous PTB using protein biomarkers, better estimation of test performance is obtained in more precisely GA-dated individuals.^33^ Since crown-rump length is more precise when determined prior to 9^0/7^ weeks’ gestation,^26^ post-validation performance metrics were calculated on a sub-cohort comprised of subjects dated by ultrasound prior to 9^0/7^-weeks’ gestation (early GA dating). Performance metrics were calculated for GA cutoffs of <37 and <34 weeks in all subjects and in nulliparous individuals. PE prevalence was calculated on the TREETOP validation cohort and was consistent with the broader U.S. population, as represented by National Vital Statistics System (NVSS) reporting^34^ and Lisonkova et al. (2013)^8^ (e.g., for PE <34 weeks: TREETOP, 1.13%; Lisonkova, 1.24%; *P*=0.78).

Guidelines^35^ set by the U.S. Preventive Services Task Force (USPSTF) for PE clinical risk factor assessment were compared to the biomarker-driven model. The TREETOP validation dataset contained most USPSTF-prescribed clinical risk factors, including prior history of preeclampsia, chronic hypertension, type 1 or type 2 diabetes, kidney disease, multi-gestational birth, elevated pre-pregnancy body mass index, instances of nulliparous births, and maternal African American heritage (attributed to socioeconomic factors). Using these variables, a classifier was developed to approximate the USPSTF risk assessment and compare its performance with biomarker-driven models using the same predefined benchmarks.

Analyses were conducted according to prespecified protocols. Laboratory personnel and statisticians were blinded to clinical data. Data linked to classifier scores were analyzed by an independent third-party statistician. Type I error was controlled by fixed sequence hypothesis testing.^36,37^ The sample size of the validation sub-cohort was shown to have 84% power at 5% alpha to detect by Fisher’s exact test an enrichment of spontaneous PTB cases above a predictor score threshold associated with two-fold increased risk.^38^ For the purpose of validating preterm PE classifiers, this sub-cohort was estimated to show >90% power at 5% alpha to detect by regression test a significant enrichment above a 75% sensitivity threshold. At such a threshold, the sub-cohort sample size was estimated to produce a 95% CI width for sensitivity of <25% and for specificity of <5%.

Unless otherwise specified, all statistical tests were 2-tailed at significance <0.05 and performed in R v3.5.1 or higher.^39^ Count differences in categorical variables were assessed using the chi-squared test, and median differences in continuous variables were assessed with the Wilcoxon test.^40^ Comparison across cohorts was performed using the Kruskal-Wallis test. Overall predictive performance was assessed via AUC with direction of effect prespecified, significance assessed with a 1-sided Wilcoxon test, and confidence intervals (CI) calculated using DeLong’s method.^41^ Clinical validity was assessed using predictive performance and statistically significant stratification of preterm PE subjects above vs below threshold scores. Following validation, sensitivity, specificity, PPV, NPV, SPR, likelihood ratios (LR+/-), and odds ratios (OR) were calculated for preterm PE (GAD <37 weeks), and early preterm PE (GAD <34 weeks) for the entire validation cohort (all parities) and for nulliparas only. Calculation of prevalence-dependent predictor performance used the prevalence of the analysed outcome in the entire TREETOP-based validation cohort, as well as the early GA dating subset.

## Results

Characteristics of the discovery, verification, and validation cohorts are summarized in **Table 1** and **Fig. 1**. Subjects delivering with preterm PE were more likely than controls to have higher BMI, prior PE, chronic hypertension, pregestational diabetes, and longer neonatal hospital stays. GABD showed no significant difference between cases and controls in any cohort.

**Table 1.**
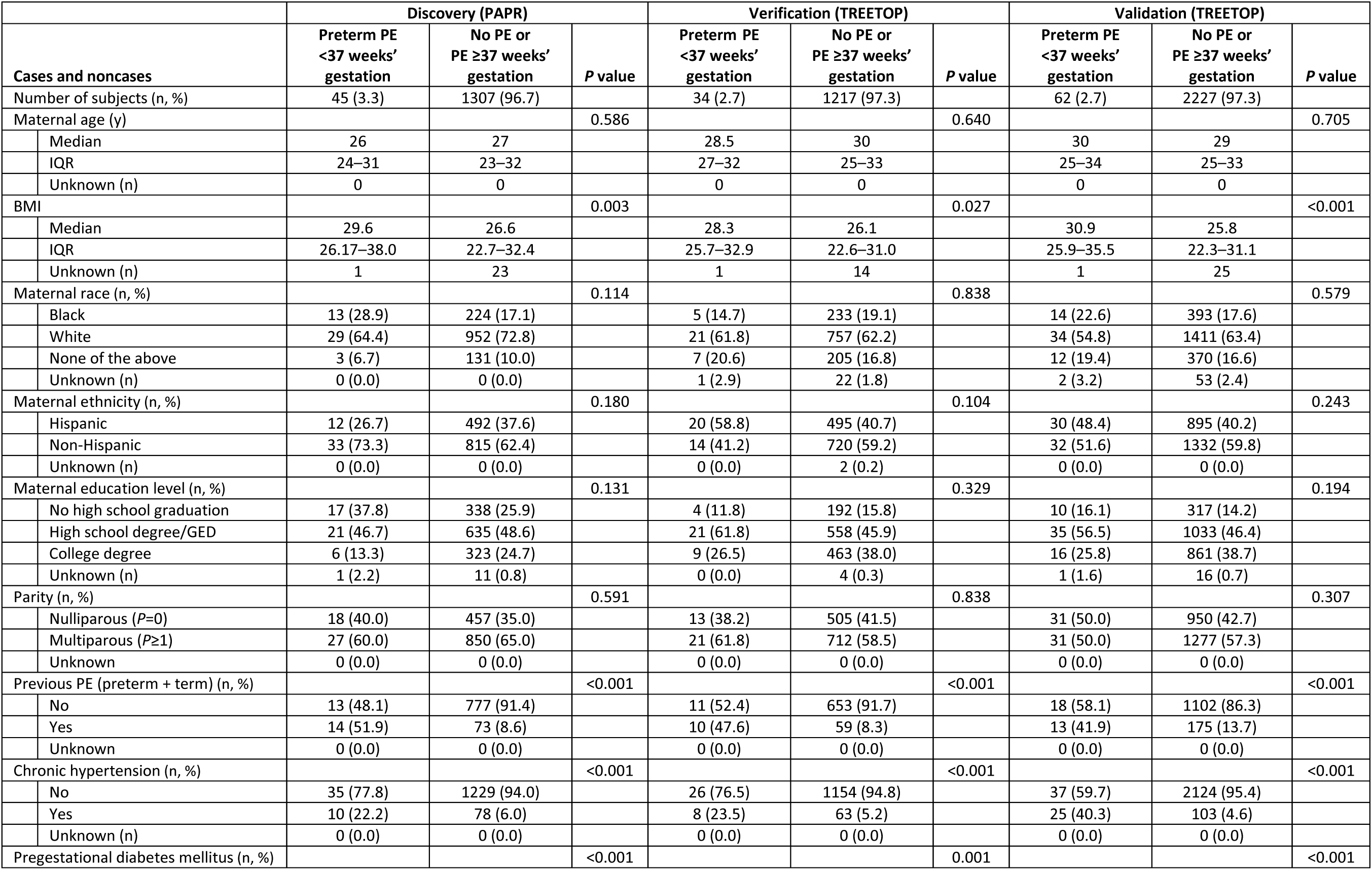
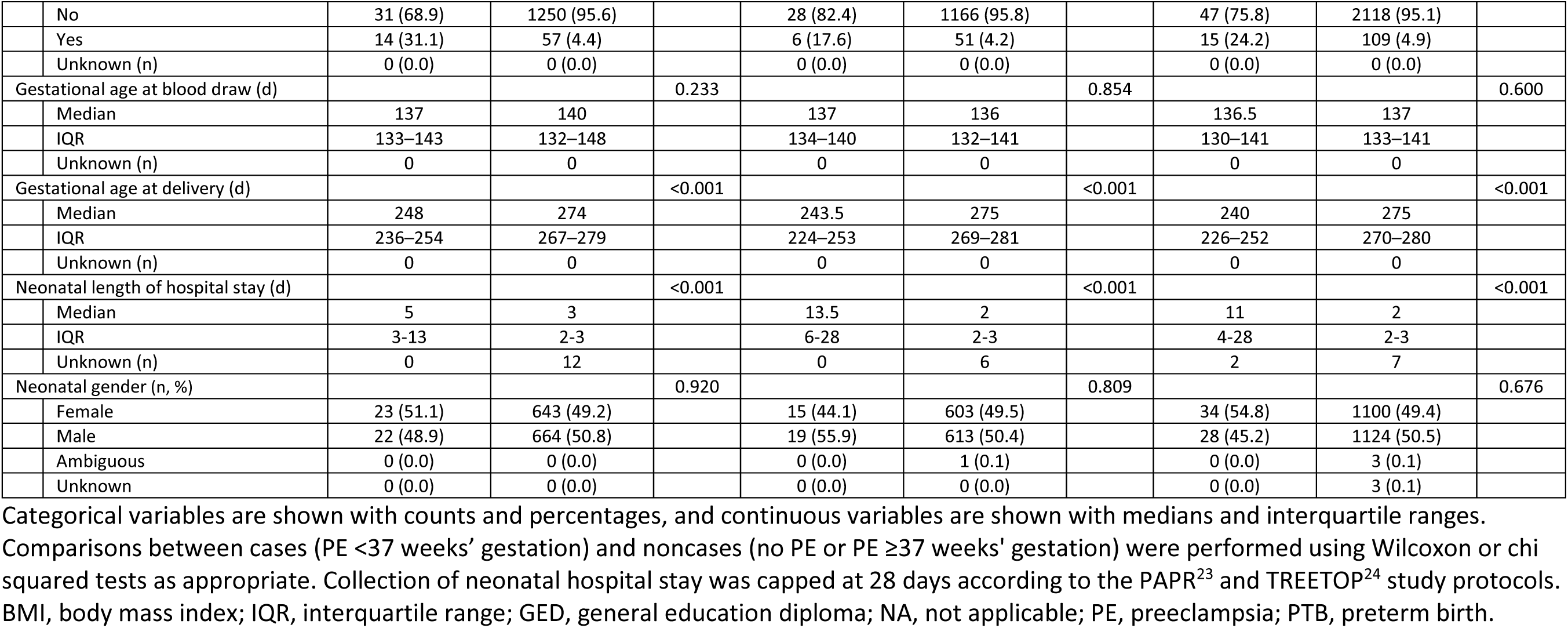
Demographic and clinical characteristics of preterm preeclampsia (PE) cases and noncases in the discovery, verification, and validation cohorts.

| Cases and noncases | Discovery (PAPR) |  |  | Verification (TREETOP) |  |  | Validation (TREETOP) |  |  |
| --- | --- | --- | --- | --- | --- | --- | --- | --- | --- |
|  | Preterm PE <37 weeks' gestation | No PE or PE ≥37 weeks' gestation | P value | Preterm PE <37 weeks' gestation | No PE or PE ≥37 weeks' gestation | P value | Preterm PE <37 weeks' gestation | No PE or PE ≥37 weeks' gestation | P value |
| Number of subjects (n, %) | 45 (3.3) | 1307 (96.7) |  | 34 (2.7) | 1217 (97.3) |  | 62 (2.7) | 2227 (97.3) |  |
| Maternal age (y) |  |  | 0.586 |  |  | 0.640 |  |  | 0.705 |
| Median | 26 | 27 |  | 28.5 | 30 |  | 30 | 29 |  |
| IQR | 24–31 | 23–32 |  | 27–32 | 25–33 |  | 25–34 | 25–33 |  |
| Unknown (n) | 0 | 0 |  | 0 | 0 |  | 0 | 0 |  |
| BMI |  |  | 0.003 |  |  | 0.027 |  |  | <0.001 |
| Median | 29.6 | 26.6 |  | 28.3 | 26.1 |  | 30.9 | 25.8 |  |
| IQR | 26.17–38.0 | 22.7–32.4 |  | 25.7–32.9 | 22.6–31.0 |  | 25.9–35.5 | 22.3–31.1 |  |
| Unknown (n) | 1 | 23 |  | 1 | 14 |  | 1 | 25 |  |
| Maternal race (n, %) |  |  | 0.114 |  |  | 0.838 |  |  | 0.579 |
| Black | 13 (28.9) | 224 (17.1) |  | 5 (14.7) | 233 (19.1) |  | 14 (22.6) | 393 (17.6) |  |
| White | 29 (64.4) | 952 (72.8) |  | 21 (61.8) | 757 (62.2) |  | 34 (54.8) | 1411 (63.4) |  |
| None of the above | 3 (6.7) | 131 (10.0) |  | 7 (20.6) | 205 (16.8) |  | 12 (19.4) | 370 (16.6) |  |
| Unknown (n) | 0 (0.0) | 0 (0.0) |  | 1 (2.9) | 22 (1.8) |  | 2 (3.2) | 53 (2.4) |  |
| Maternal ethnicity (n, %) |  |  | 0.180 |  |  | 0.104 |  |  | 0.243 |
| Hispanic | 12 (26.7) | 492 (37.6) |  | 20 (58.8) | 495 (40.7) |  | 30 (48.4) | 895 (40.2) |  |
| Non-Hispanic | 33 (73.3) | 815 (62.4) |  | 14 (41.2) | 720 (59.2) |  | 32 (51.6) | 1332 (59.8) |  |
| Unknown (n) | 0 (0.0) | 0 (0.0) |  | 0 (0.0) | 2 (0.2) |  | 0 (0.0) | 0 (0.0) |  |
| Maternal education level (n, %) |  |  | 0.131 |  |  | 0.329 |  |  | 0.194 |
| No high school graduation | 17 (37.8) | 338 (25.9) |  | 4 (11.8) | 192 (15.8) |  | 10 (16.1) | 317 (14.2) |  |
| High school degree/GED | 21 (46.7) | 635 (48.6) |  | 21 (61.8) | 558 (45.9) |  | 35 (56.5) | 1033 (46.4) |  |
| College degree | 6 (13.3) | 323 (24.7) |  | 9 (26.5) | 463 (38.0) |  | 16 (25.8) | 861 (38.7) |  |
| Unknown (n) | 1 (2.2) | 11 (0.8) |  | 0 (0.0) | 4 (0.3) |  | 1 (1.6) | 16 (0.7) |  |
| Parity (n, %) |  |  | 0.591 |  |  | 0.838 |  |  | 0.307 |
| Nulliparous (P=0) | 18 (40.0) | 457 (35.0) |  | 13 (38.2) | 505 (41.5) |  | 31 (50.0) | 950 (42.7) |  |
| Multiparous (P≥1) | 27 (60.0) | 850 (65.0) |  | 21 (61.8) | 712 (58.5) |  | 31 (50.0) | 1277 (57.3) |  |
| Unknown | 0 (0.0) | 0 (0.0) |  | 0 (0.0) | 0 (0.0) |  | 0 (0.0) | 0 (0.0) |  |
| Previous PE (preterm + term) (n, %) |  |  | <0.001 |  |  | <0.001 |  |  | <0.001 |
| No | 13 (48.1) | 777 (91.4) |  | 11 (52.4) | 653 (91.7) |  | 18 (58.1) | 1102 (86.3) |  |
| Yes | 14 (51.9) | 73 (8.6) |  | 10 (47.6) | 59 (8.3) |  | 13 (41.9) | 175 (13.7) |  |
| Unknown | 0 (0.0) | 0 (0.0) |  | 0 (0.0) | 0 (0.0) |  | 0 (0.0) | 0 (0.0) |  |
| Chronic hypertension (n, %) |  |  | <0.001 |  |  | <0.001 |  |  | <0.001 |
| No | 35 (77.8) | 1229 (94.0) |  | 26 (76.5) | 1154 (94.8) |  | 37 (59.7) | 2124 (95.4) |  |
| Yes | 10 (22.2) | 78 (6.0) |  | 8 (23.5) | 63 (5.2) |  | 25 (40.3) | 103 (4.6) |  |
| Unknown (n) | 0 (0.0) | 0 (0.0) |  | 0 (0.0) | 0 (0.0) |  | 0 (0.0) | 0 (0.0) |  |
| Pregestational diabetes mellitus (n, %) |  |  | <0.001 |  |  | 0.001 |  |  | <0.001 |

|  |  |  |  |  |  |  |  |  |  |  |
| --- | --- | --- | --- | --- | --- | --- | --- | --- | --- | --- |
|  | No | 31 (68.9) | 1250 (95.6) |  | 28 (82.4) | 1166 (95.8) |  | 47 (75.8) | 2118 (95.1) |  |
|  | Yes | 14 (31.1) | 57 (4.4) |  | 6 (17.6) | 51 (4.2) |  | 15 (24.2) | 109 (4.9) |  |
|  | Unknown (n) | 0 (0.0) | 0 (0.0) |  | 0 (0.0) | 0 (0.0) |  | 0 (0.0) | 0 (0.0) |  |
| Gestational age at blood draw (d) |  |  |  | 0.233 |  |  | 0.854 |  |  | 0.600 |
|  | Median | 137 | 140 |  | 137 | 136 |  | 136.5 | 137 |  |
|  | IQR | 133–143 | 132–148 |  | 134–140 | 132–141 |  | 130–141 | 133–141 |  |
|  | Unknown (n) | 0 | 0 |  | 0 | 0 |  | 0 | 0 |  |
| Gestational age at delivery (d) |  |  |  | <0.001 |  |  | <0.001 |  |  | <0.001 |
|  | Median | 248 | 274 |  | 243.5 | 275 |  | 240 | 275 |  |
|  | IQR | 236–254 | 267–279 |  | 224–253 | 269–281 |  | 226–252 | 270–280 |  |
|  | Unknown (n) | 0 | 0 |  | 0 | 0 |  | 0 | 0 |  |
| Neonatal length of hospital stay (d) |  |  |  | <0.001 |  |  | <0.001 |  |  | <0.001 |
|  | Median | 5 | 3 |  | 13.5 | 2 |  | 11 | 2 |  |
|  | IQR | 3–13 | 2–3 |  | 6–28 | 2–3 |  | 4–28 | 2–3 |  |
|  | Unknown (n) | 0 | 12 |  | 0 | 6 |  | 2 | 7 |  |
| Neonatal gender (n, %) |  |  |  | 0.920 |  |  | 0.809 |  |  | 0.676 |
|  | Female | 23 (51.1) | 643 (49.2) |  | 15 (44.1) | 603 (49.5) |  | 34 (54.8) | 1100 (49.4) |  |
|  | Male | 22 (48.9) | 664 (50.8) |  | 19 (55.9) | 613 (50.4) |  | 28 (45.2) | 1124 (50.5) |  |
|  | Ambiguous | 0 (0.0) | 0 (0.0) |  | 0 (0.0) | 1 (0.1) |  | 0 (0.0) | 3 (0.1) |  |
|  | Unknown | 0 (0.0) | 0 (0.0) |  | 0 (0.0) | 0 (0.0) |  | 0 (0.0) | 3 (0.1) |  |

Categorical variables are shown with counts and percentages, and continuous variables are shown with medians and interquartile ranges. Comparisons between cases (PE <37 weeks' gestation) and noncases (no PE or PE ≥37 weeks' gestation) were performed using Wilcoxon or chi squared tests as appropriate. Collection of neonatal hospital stay was capped at 28 days according to the PAPR<sup>23</sup> and TREETOP<sup>24</sup> study protocols. BMI, body mass index; IQR, interquartile range; GED, general education diploma; NA, not applicable; PE, preeclampsia; PTB, preterm birth.

No significant differences were seen across cohorts for chronic hypertension and pregestational diabetes, but prior PE was increased among validation cohort controls (**Appendix 1**). There were also significant differences between cohorts in race, ethnicity, parity, educational attainment, maternal age, BMI, and GAD. Despite both case/control and cohort differences, regression analysis showed that maternal age and BMI did not contribute independently to preterm PE risk. The significant difference in GABD reflected the broader collection window in the discovery cohort.

Nine of nine classifiers ascertained in discovery showed acceptable predictive performance in verification for risk of preterm PE with GAD <37 weeks. In validation, all nine exhibited significant AUCs (0.72-0.78) for preterm PE risk prediction, significant risk stratification at three threshold scores, and significant Pearson correlations between predictor score and GAD among individuals with PE (**Appendix 2**). ClinRisk3 alone exhibited an AUC of 0.68 for preterm PE, falling below the lower 95% CI of most of the validated combined classifiers. **Fig. 2** shows ROC curves for the predictor with the largest AUC (0.78), ClinRisk3 + inhibin beta c chain/SHBG (ClinRisk3+INHBC/SHBG), in the full validation cohort for (A) all parity and (B) nulliparas.

**Fig. 2.**
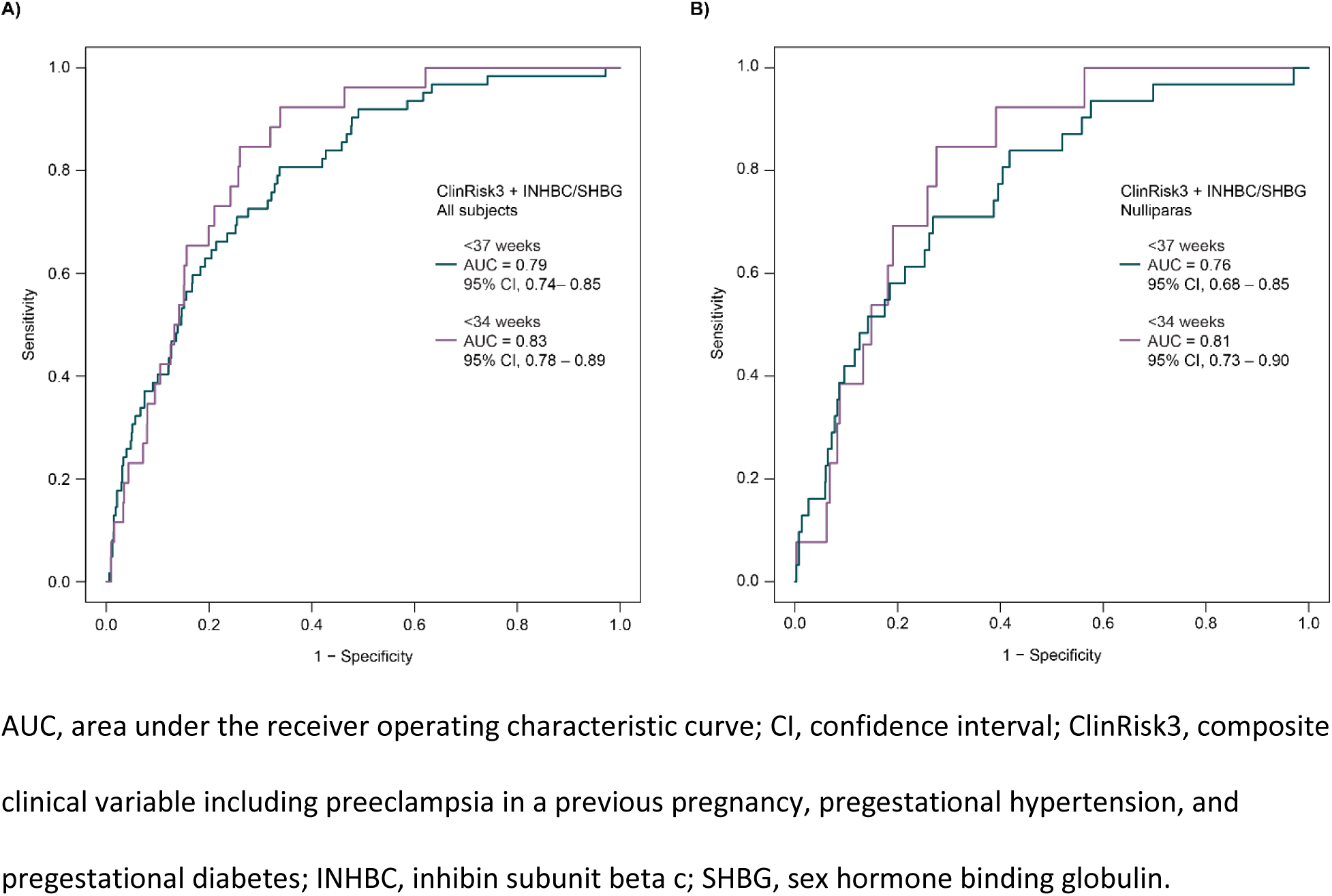
Receiver operating characteristic performance for the preterm preeclampsia (PE) classifier ClinRisk3+INHBC/SHBG in the full validation cohort. The plots graph sensitivity (true positive rate) vs 1-specificity (false positive rate) for preterm PE cases with gestational age at delivery <37 and <34 weeks among (A) all subjects and (B) nulliparous subjects only.

For prediction of preterm PE <37 weeks’ gestation in gravidas with any parity or nulliparas, **Table 2** details ClinRisk3+INHBC/SHBG performance for validation cohort subjects with early GA dating, and **Appendix 3** shows its performance in the full validation cohort. In the early GA dating sub-cohort, setting the ClinRisk3+INHBC/SHBG cutoff to achieve a specificity of at least 90%, the sensitivity for prediction of preterm PE with GAD <37 weeks for all parities was 51.7%, with a SPR 11.1%, PPV 13.0%, NPV 98.5%, and OR 9.93; the performance for nulliparas was extremely similar, with the exception of a lower PPV (6.9%) and higher NPV (99.8%). We also applied USPSTF clinical practice guidelines^35^ to our validation cohort – either the entire validation cohort (**Appendix 3**) or the sub-cohort with early GA dating (**Table 2**). In these comparisons, predictive performance for the combination of ClinRisk3 and INHBC/SHBG was superior to that of USPSTF guidelines.

**Table 2.** Predictive performance of ClinRisk3+INHBC/SHBG and a U.S. Preventive Services Task Force clinical risk factor-based classifier (representing standard care) for risk of preeclampsia with delivery <37 weeks’ gestation in the early gestational age dating-restricted population.

| Tested population | PE <37 weeks' GA cases/ total subjects (%) | Predictor | AUC (95% CI) | Sensitivity | Specificity | SPR | PPV | NPV | LR+ | LR- | OR |
| --- | --- | --- | --- | --- | --- | --- | --- | --- | --- | --- | --- |
| Early GA dating<br>All parity | 29/912 (3.2%) | ClinRisk3 + INHBC/SHBG | 0.82 (0.75 – 0.89) | 51.7% | 90.3% | 11.1% | 13.0% | 98.5% | 5.31 | 0.53 | 9.93 |
|  |  | USPSTF guidelines (standard of care) | 0.69 (0.61 – 0.76) | 79.3% | 57.8% | 43.4% | 5.0% | 99.0% | 1.88 | 0.36 | 5.24 |
| Early GA dating<br>Nulliparas | 15/376 (4.0%) | ClinRisk3 + INHBC/SHBG | 0.81 (0.71 – 0.92) | 53.3% | 90.0% | 11.7% | 6.9% | 99.8% | 5.35 | 0.52 | 10.32 |
|  |  | USPSTF guidelines (standard of care) | 0.70 (0.61 – 0.80) | 86.7% | 54.3% | 47.3% | 2.6% | 99.1% | 1.90 | 0.25 | 7.72 |

AUC, area under the receiver operating characteristic curve; CI, confidence interval; ClinRisk3, composite clinical risk variable including preeclampsia in a previous pregnancy, pregestational hypertension, and pregestational diabetes; GA, gestational age; INHBC, inhibin subunit beta C chain; NPV, negative predictive value; LR+/-, positive and negative likelihood ratios; OR, odds ratio; PPV, positive predictive value; SHBG, sex hormone-binding globulin; SPR, screen positive rate; USPSTF, U.S. Preventive Services Task Force.

## Discussion

Nine classifier models predictive of preterm PE risk were validated, with ClinRisk3+INHBC/SHBG showing the best overall performance. Requiring simplicity (one or two bivariate protein biomarkers and a composite clinical variable) and using large, diverse, and temporally divergent cohorts mitigated overfitting and enabled validation in an independent population. Three clinical features that differed significantly between cases and controls in the discovery cohort (prior PE, pre-existing hypertension, and pregestational diabetes) comprised the composite clinical variable. A fourth, BMI, was excluded as it did not contribute independently to preterm PE risk, perhaps due to strong associations between BMI and both diabetes and SHBG.^42,43^ Emphasizing strong predictive performance in both nulliparous and multiparous pregnancies during discovery ensures broad applicability. In the two source cohorts, Black (17.8-19.0%) and Hispanic (37.3-41.2%) subjects were represented at proportions higher than those in the general U.S. birthing population (14.1-15.2% Black; 23.7-24.2% Hispanic),^44,45^ ensuring generalizability.

Inhibin and activin, members of the transforming growth factor beta (TGFβ) family, are dysregulated in pregnancy disorders. Elevated maternal serum Inhibin A was first reported as a biomarker of fetal aneuploidy^46^ but is also associated with PE,^47,48^ including preclinical PE.^49^ However, Inhibin A, alone or in combination with other aneuploidy and fetal anomaly biomarkers, demonstrates insufficient predictive performance for clinical use.^50,51^ INHBC, a less well-characterized inhibin family member comprised of activin C homodimers, was recently reported to function through activin receptor-like kinase 7 (ALK7), which participates in angiogenesis, cytokine activity, hormone signaling, and embryonic development.^52^ Expressed in villous and extravillous trophoblasts, ALK7 is dysregulated in placentas from individuals with PE^53^ and is a mediator of hypoxia-induced impairment trophoblast invasion.^54^ A potential role for INHBC in extravillous trophoblast development and function may help explain its predictive value for preterm PE (**Fig. 3**).

**Fig. 3.**
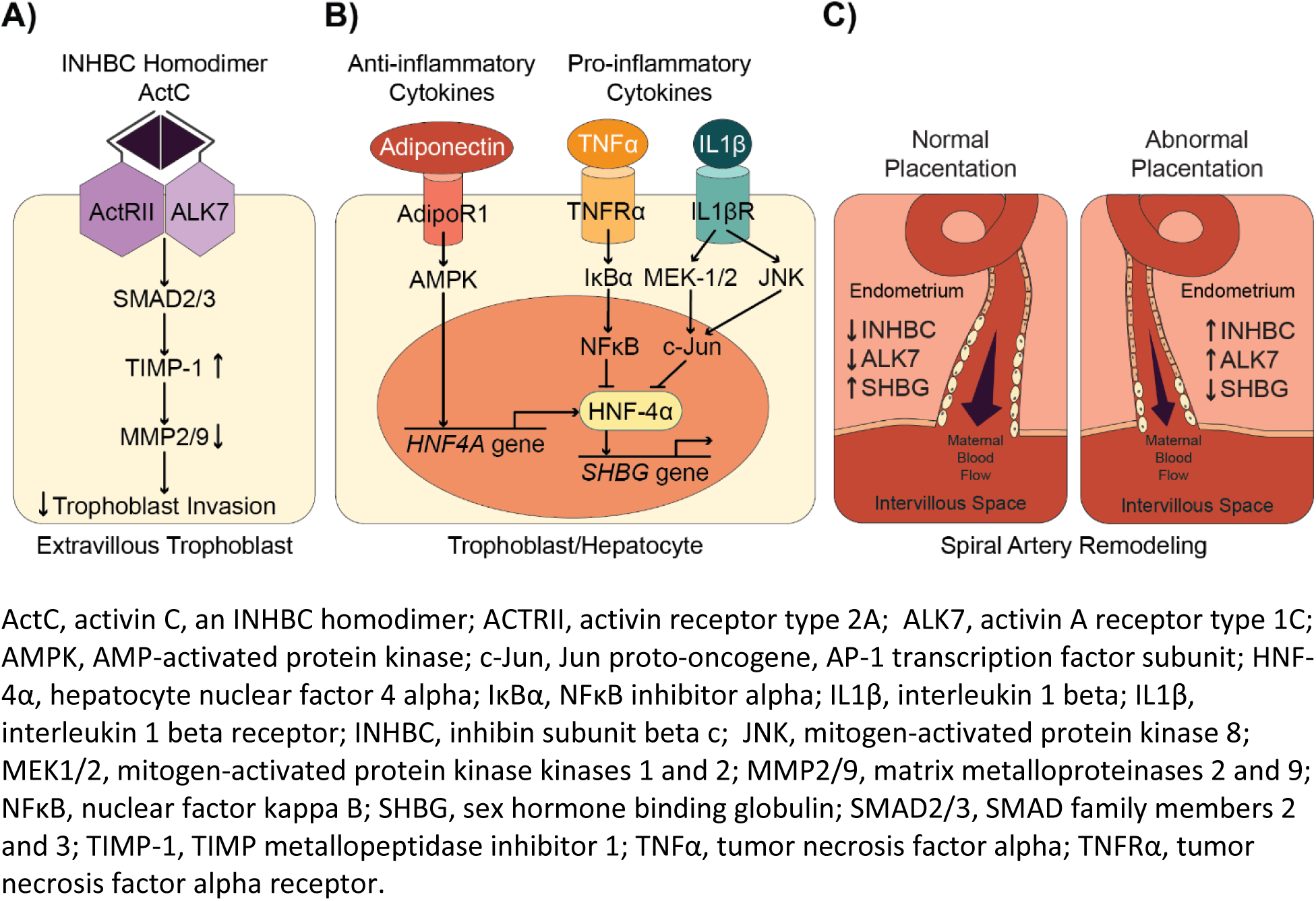
Potential roles of INHBC and SHBG in placentation. (A) Activin C, a homodimer of INHBC, signals through the ALK7 receptor, which is itself dysregulated in preeclampsia. (B) SHBG expression has been demonstrated to be downregulated by pro-inflammatory cytokines in hepatocytes, and its extra-hepatic expression has been shown to include trophoblasts. (C) A model illustrating the association of INHBC, ALK7 and SHBG with abnormal placentation and aberrant spiral artery remodeling.

SHBG regulates biologically active free steroid hormone levels,^55^ increases up to 10-fold in the maternal circulation during pregnancy, and is expressed by placental trophoblasts.^55,56^ SHBG transcription is suppressed by pro-inflammatory cytokines, such as tumor necrosis factor-alpha (TNF-α),^57^ which is relevant because inflammation, along with increased placental^58^ and circulating^59^ TNF-α, is associated with PE (**Fig. 3**). Dysregulation of maternal circulating SHBG in PE has been reported by some^60^ and disputed by others^61–65^, perhaps due to variability in PE phenotype and GABD among studies. Even in studies associating SHBG with PE, its predictive performance is poor.^60^ In non-pregnant adults, low serum SHBG has been associated with insulin resistance,^66^ Type 2 diabetes,^67^ and endothelial dysfunction.^68^ In one study, gravidas with low first-trimester SHBG and PlGF were at highest risk of developing PE, leading authors to conclude that dysregulated angiogenesis and insulin resistance exerted additive effects on PE risk.^66^ Similarly, these results suggest that SHBG is an independent PE risk predictor due to potential effects on insulin resistance and endothelial function, and predictive performance improves markedly when combined with an angiogenic factor such as INHBC.

Since the validated ClinRisk3+INHBC/SHBG predictor makes early risk assessment possible for preterm PE (<37 weeks) in a given pregnancy for any parity, it can enable individualized clinical management. Individuals at higher risk can be assigned to increased surveillance (e.g., more frequent antenatal visits, home blood pressure monitoring) for early detection of PE and associated complications, along with nutritional modification, exercise coaching, and other interventions,^69^ while those at lower risk can safely avoid unnecessary interventions. Determining whether our predictors will be useful for decision-making regarding LDASA prophylaxis requires more study, to establish LDASA efficacy among patients for whom guideline criteria and the predictors produce discordant risk assessments.

ClinRisk3+INHBC/SHBG provided excellent stratification of subjects at elevated risk for early-onset preterm PE (diagnosis <34 weeks’ GA), an outcome with a large clinical burden and unmet need. ACOG recommends delivery for PE diagnosed ≥34 weeks’ GA and expectant management with close observation for those diagnosed earlier.^7^ Clark et al. (2023)^70^ recently recommended changes to classification of different PE subtypes to better reflect the type and degree of associated organ dysfunction. We acknowledge that, if adopted, such recommendations may alter clinical decision-making relevant to timing of delivery for PE. In addition, new prevention or treatment strategies may be developed. Any of these developments may require us to reassess predictor performance in the appropriate contexts.

Clinical practice guidelines include expectations for test performance, including high PPV, to limit overtreatment.^7^ However, PPV is impacted by a condition’s prevalence. This study validated predictive algorithms for preterm PE, a clinically important yet low-prevalence PE phenotype, in a cohort with a PE rate similar to the overall U.S. prevalence (2.7% preterm PE, 7.7% all PE). This contrasts with a recent RNA biomarker study^71^ that interrogated PE at any GA in a high-risk population (PE rate of 13.6%), which reported a sensitivity of 75.0% and a PPV of 32.3%. We chose to establish a threshold at approximately a 10% false positive rate (FPR), as this is a common practice that helps to avoid overtreatment. It is also common to establish a threshold that results in 10% of a population being screen positive. As our 10% thresholds result in approximately an 11-12% SPR, the performance estimates provided here apply generally for both approaches.

Importantly, according to the current standard of care, PE risk is assessed using a range of clinical algorithms. Specifically, in a large Danish mixed parity cohort, National Institute of Health and Care Excellence (NICE) guidelines^28^ had a sensitivity of 47%, PPV of 5.4%, and NPV of 99.6% at a SPR of 8.1%, while American College of Obstetrics and Gynecology (ACOG) guidelines^7^ had a sensitivity of 60.5%, PPV of 5.6%, and NPV of 99.7% at a SPR of 18.2% for preterm preeclampsia <37 weeks’ GA.^72^ In our validation cohort, we applied USPSTF clinical practice guidelines, which yielded a sensitivity of 79.3%, PPV of 5.0%, and NPV of 99.0% at a SPR of 43.4% for preterm preeclampsia <37 weeks’ GA. The Fetal Medicine Foundation’s (FMF) recommended a more involved screening algorithm^73^, including maternal risk factors, maternal mean arterial pressure, the uterine artery pulsatility index, and serum PLGF, which has a reported 79.9% sensitivity, 4.4% PPV, and 99.8% NPV at a SPR of 14.7% for all parities, and 79.0% sensitivity, 3.8% PPV, and 99.8% NPV at a SPR of 19.2% for nulliparas, for preterm PE.^74^

Consisting of straightforward measurement of 2 proteins in a single blood sample combined with easily ascertained clinical factors, the performance of ClinRisk3+INHBC/SHBG in the total validation cohort (sensitivity 51.7%, PPV of 13.0%, and NPV of 98.5% at a SPR of 11.1%) and specifically in nulliparas (sensitivity 53.3%, PPV of 6.9%, and NPV of 99.8% at a SPR of 11.7%) is superior to standard-of-care clinical risk assessment approaches endorsed by NICE, ACOG, and USPSTF, and also compares favorably with the more burdensome approach endorsed by the FMF, which is comprised of not only clinical factors and a protein biomarker, but also uterine artery Doppler measurement, which is unfeasible for universal screening. ClinRisk3+INHBC/SHBG and the other predictors validated in this study can be used to select high-risk pregnancies for interventional studies of prophylactic or therapeutic agents. Determining whether preventive or therapeutic interventions alter biomarker levels in maternal serum is also of interest. Clinical application of a screening test is a two-step process: The first step, reported here, is validation of the biomarkers’ predictive abilities. The second, demonstrating clinical utility – i.e., improved outcomes, cost effectiveness, and ease of implementation – requires additional work.

Strengths of the study include the large, diverse, and temporally distinct datasets used for discovery, verification, and validation. The composite clinical variable uses readily ascertained factors, and the biomarkers were prefiltered for excellent performance in ambient temperature sample transport conditions. These properties support practicality for clinical use. Limitations include the modest number of early preterm PE cases due to the low prevalence of the condition and a mid-trimester blood draw window (18^0/7^-22^6/7^ weeks’ GA) that falls within the guideline-recommended timeframe for LDASA initiation (12-28 week’s GA, optimally <16 weeks’ GA).^7^

In summary, nine mid-trimester preterm PE risk predictors, designed to assess both proteomic biomarkers and clinical risk factors from 18-20 weeks’ gestation, demonstrated predictive ability superior to that of clinical factors alone. ClinRisk3+INHBC/SHBG includes biomarkers with functional relevance to pregnancy. All show promise for detecting risk of PE <34 weeks’ GA and identifying pre-symptomatic individuals who can benefit from increased surveillance, LDASA prophylaxis, and future therapies.

## Data Availability

All data produced in the present study are available upon reasonable request to the authors.

## Acknowledgments

Study funding was provided by Sera Prognostics, Inc. Jennifer Logan, PhD, an employee of Sera Prognostics, Inc., contributed to the writing of this article. Babak Shahbaba, PhD (University of California Irvine), conducted independent validation hypothesis testing. The authors wish to acknowledge the following co-investigators on the original PAPR and/or TREETOP studies: Kim A. Boggess, MD (University of North Carolina School of Medicine); Dean V. Coonrod, MD (University of Arizona College of Medicine); Larry M. Cousins, MD (San Diego Perinatal Center; Rady Children’s Specialists of San Diego); Amy H. Crockett, MD (University of South Carolina School of Medicine Greenville and Prisma Health-Upstate); M. Sean Esplin, MD (Intermountain Healthcare); William A. Grobman, MD, MBA (The Ohio State University Wexner Medical Center); David M. Haas, MD (Indiana University School of Medicine); Angela F. Hawk, MD (Regional Obstetrical Consultants); Jay D. Iams, MD (The Ohio State University Wexner Medical Center); Carol A. Major, MD (University of California Irvine School of Medicine); Scott A. Sullivan, MD (Medical University of South Carolina); and Sarahn M. Wheeler, MD (Duke University School of Medicine). The authors also thank the study coordinators and research personnel at the study sites, the clinical study participants, and the Sera Prognostics, Inc. research and development, clinical laboratory and clinical operations teams.

## Supplemental Digital Content

## Appendix 1.

Comparison of demographic and clinical characteristics across the discovery, verification, and validation cohorts.

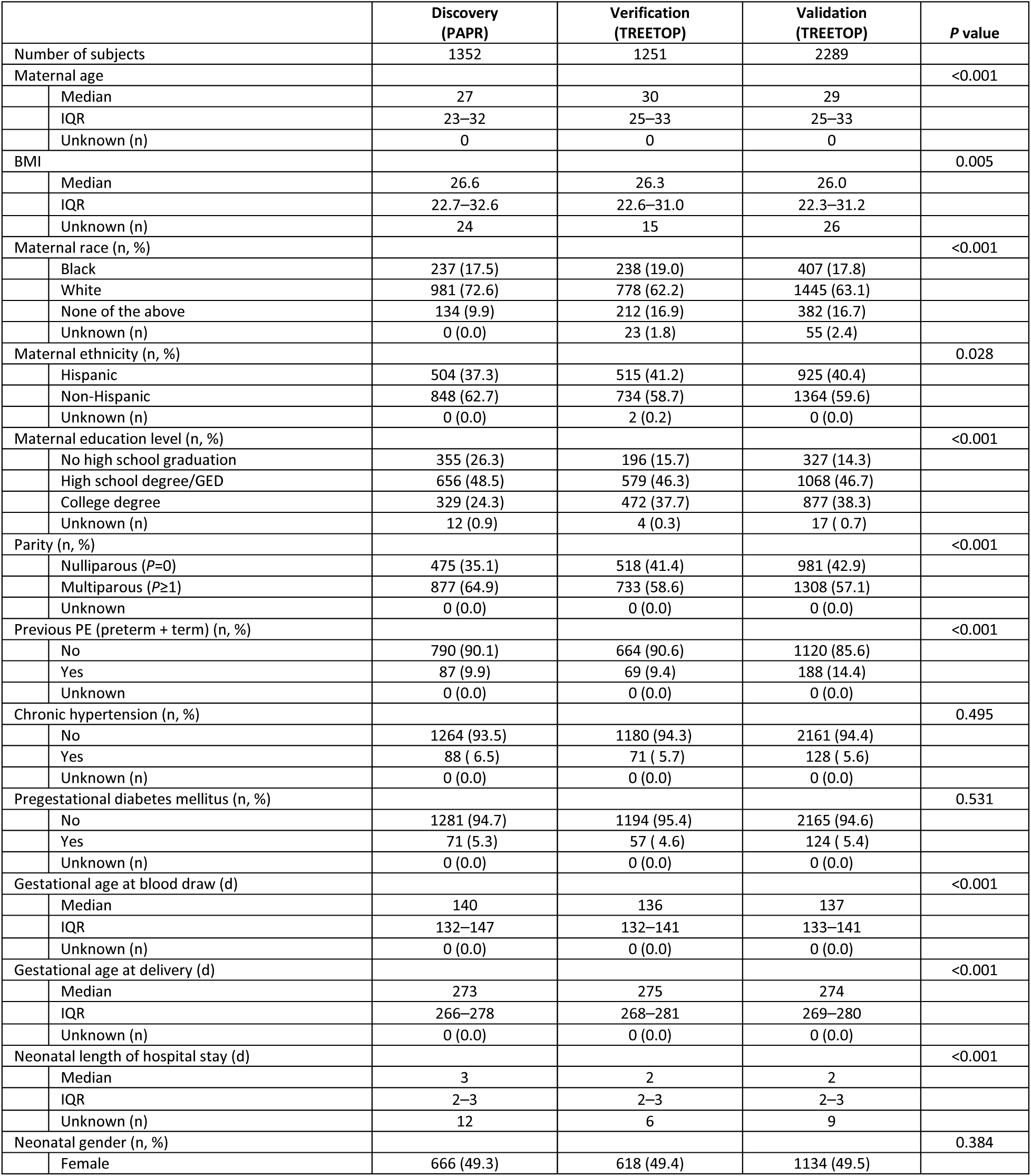
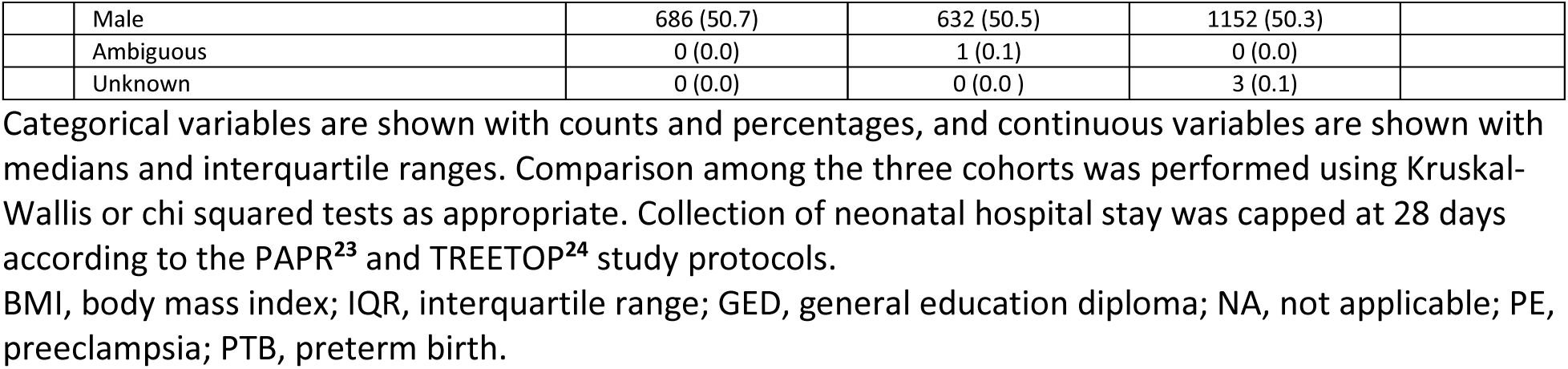

## Appendix 2.

Validation performance of nine candidate preeclampsia classifier models. Score thresholds are specific to each classifier and differentiate subjects predicted to be at higher risk for preterm preeclampsia, defined as preeclampsia <37 weeks’ gestation, from those predicted to be at lower risk. Three score thresholds distinguishing higher from lower risk were validated for each classifier. The linear relationship between the gestational age at delivery and classifier prediction was assessed by the Pearson correlation coefficient. Validation performance is reported across the nine classifiers without protein specific batch calibration.

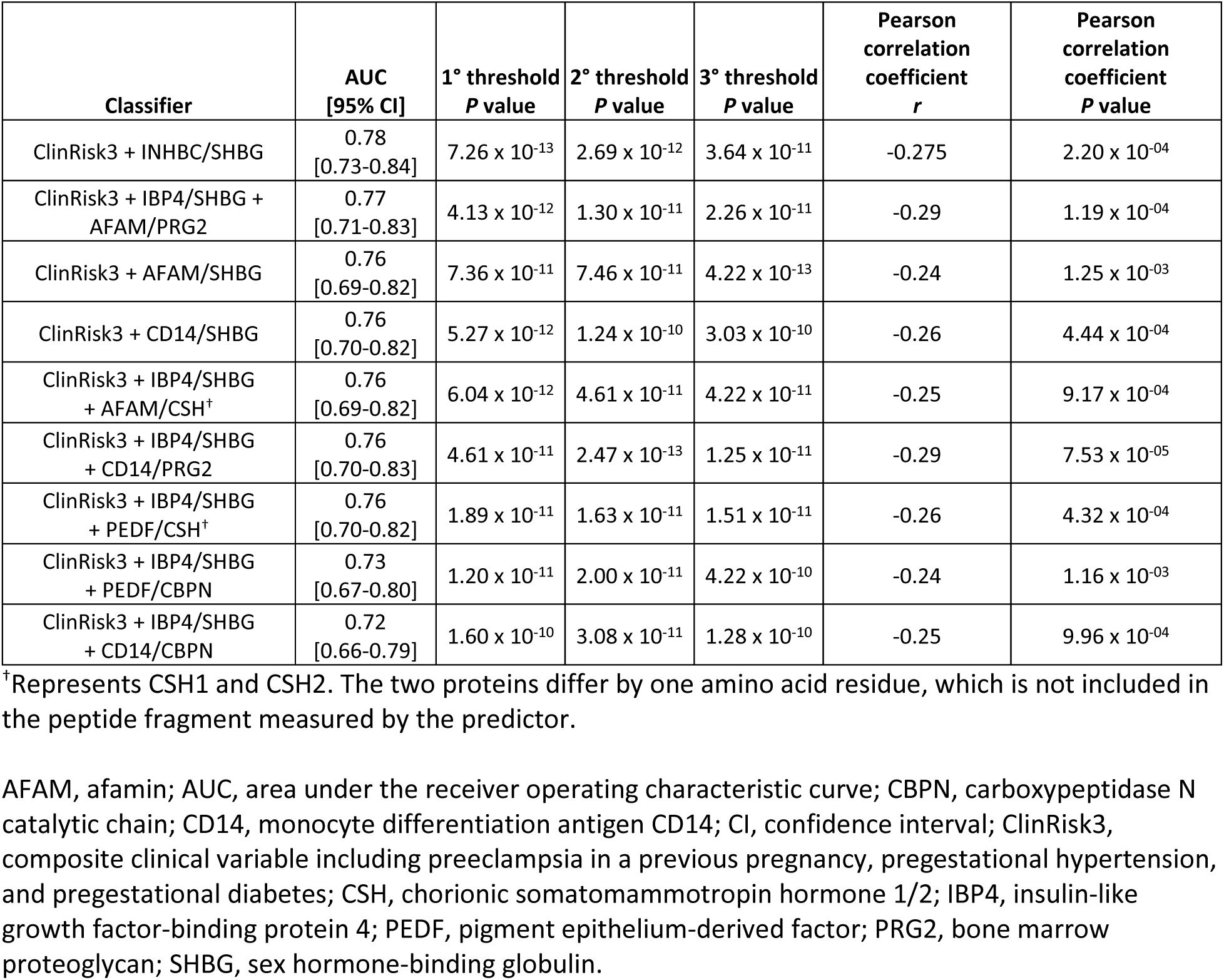

## Appendix 3.

Predictive performance of the ClinRisk3+INHBC/SHBG classifier model and a U.S. Preventive Services Task Force^35^ clinical risk factor-based classifier (representing standard care) for preterm preeclampsia risk in the full validation cohort.

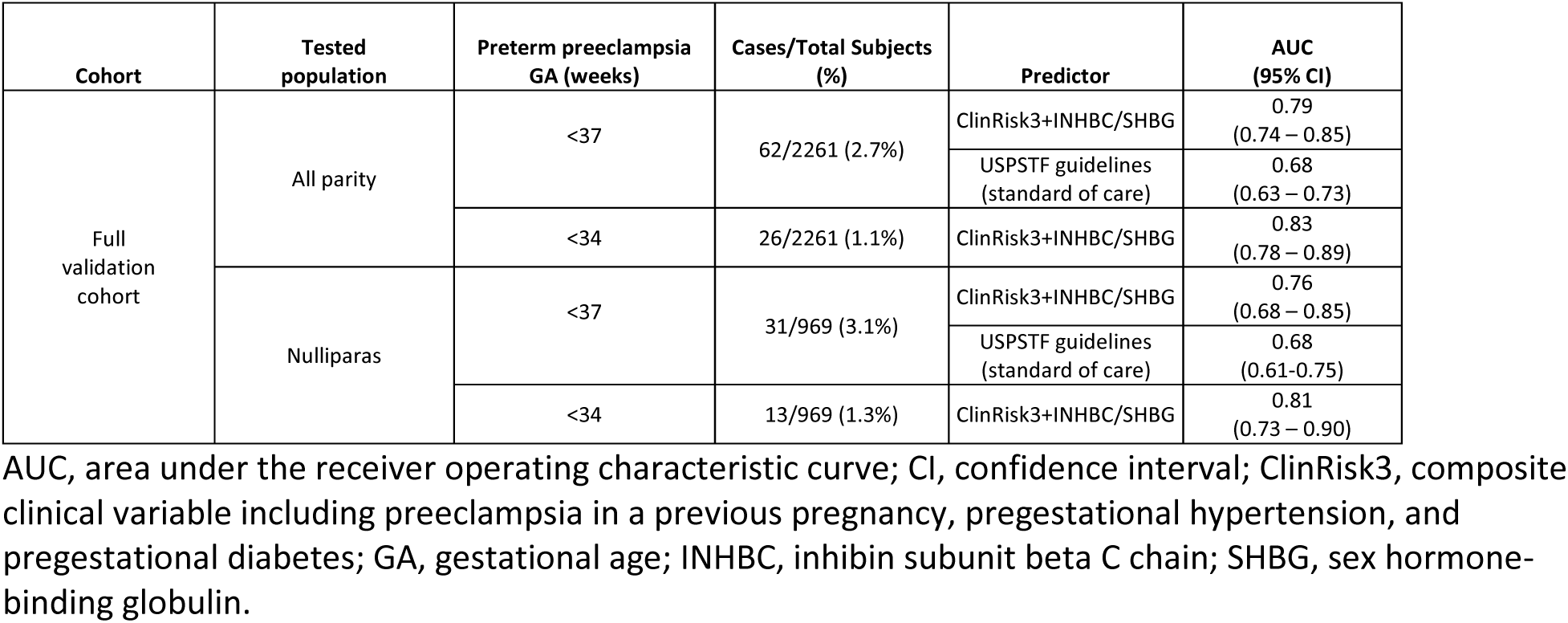

